# Mortality and Mode of Dialysis

**DOI:** 10.1101/2023.02.14.23285929

**Authors:** Subhash Chander, Sindhu Lohana, FNU Sadarat, Roopa Kumari

**Affiliations:** Department of Medicine, Mount Sinai Beth Israel, NY, New York, USA; Department of Medicine AGA khan University Hospital, Karachi, Pakistan; Department of Medicine, University at Buffalo; Department of Pathology, Mount Sinai Morningside and West, New York, USA

**Author notes:** **Corresponding Author:** Subhash Chander, MBBS., MPH, **Address:** 281 1st Ave, New York, NY 10003, **Email:**.

**Keywords:** Hemodialysis, Peritoneal dialysis, CRRT, Continuous Renal Replacement Therapy

## Abstract

**Background:** Dialysis is an intervention used chiefly to cover poorly functioning kidneys’ ultrafiltration and diffusion functions. Dialysis can be performed using three major approaches: peritoneal dialysis, hemodialysis, and continuous renal replacement therapy, which have varying degrees of efficiency. We sought to compare the mortality rates among patients receiving different dialysis modalities.

**Study Design:** Statistical Review and Meta-Analysis.

**Setting & Population:** The investigation was conducted according to Preferred Reporting Items for Systematic Reviews and Meta-Analyses (PRISMA). Data sources were drawn from Google Scholar, ResearchGate, PubMed (MEDLINE), Cochrane Library, and Embase databases. Eligible patients had to be ones presenting with acute or chronic kidney failure who required assistance with kidney function.

**Selection Criteria for Studies:** The eligibility criteria included studies with participants requiring dialysis and comparing two of the three dialysis modalities that provided outcomes on mortality rates. Inclusion criteria (underlying disease, chronic or acute kidney disease, presenting signs, age categories, subject consent, etc.), the type of dialysis modalities under investigation, and the mortality rates (% per modality group).

**Index Texts:** ‘mortalities, fatalities, dialysis, hemodialysis, continuous renal replacement therapy (CRRT), Peritoneal dialysis, and comparative.’

**Outcomes:** We sought to explore the studies’ findings by comparing the mortality rates among the three broad categories of dialysis. Therefore, we aimed to compare mortality rates in CRRT vs. Hemodialysis, mortality Rates in CRRT vs. PD, and mortality rates in patients with PD vs. HD.

**Results:** Fifteen studies were narrowed down from the study search and were placed into three categories: PD vs. CRRT (4), PD vs. HD (5), and CRRT vs. HD (6). For the three outcomes, none recorded statistically significant differences in mortality rates between the patient categories (p= 0.92, 0.009, 0.22).

**Conclusion:** Like other interventions for patients with chronic kidney disease, dialysis is associated with detrimental effects caused by the inflammatory response and worsening progression of the condition. Mortality is a common complication among dialysis patients, independent of modality, based on the lack of statistically significant differences between the three groups.

## Introduction

Dialysis is a medical intervention involving the removal of waste and excess water from the blood, mimicking kidney function. These are for patients undergoing dialysis and those whose kidneys have entirely lost their function, especially in chronic renal failure. Chronic kidney disease affects over 10% of the global population and is usually positively diagnosed when the glomerular filtration rate is less than 60 ml/min.^1^ Additionally, the patients were clinically diagnosed with albuminuria, hematuria, and cystic kidney disease. The risk of chronic kidney disease depends on many influential factors, including socioeconomic status. Individuals from low- and middle-income groups are more affected than those from low-income societies. Underlying conditions such as hypertension and glomerulonephritis increase the susceptibility of individuals to chronic kidney failure.^1^. Environmental factors such as air pollution, especially in Asian nations, increase the risk of developing chronic kidney failure. Lastly, chronic kidney disease has been associated with genetic factors hence the familiar risk of developing the condition.^1^ For a positive diagnosis of chronic kidney disease, abnormal kidney function should be positively diagnosed for over three months.

Additionally, as mentioned earlier, individuals with chronic kidney disease should present with markers of kidney damage, such as albuminuria, increased serum creatinine levels, and abnormal urinary components, such as crystal-cast electrolytes and other histological components. Pathognomonic indicators of chronic kidney failure include a reduction in the glomerular filtration rate and an increase in the albumin to creatinine ratio, usually indicating the potential worsening of the condition with possible mortality. Patients with chronic kidney disease are classified based on creatinine clearance and glomerular filtration rates, thereby allowing for better management of patients through decisions on the degree of monitoring required by the patient. Furthermore, the foundation recommends routine screening of individuals over the age of 60 or those with a medical history of cardiovascular disturbances.^2^ Patients with systemic diseases, such as kidney stones and autoimmune disorders, should also be considered for routine screening of chronic kidney disease markers.^2^ Kidney Disease: Improving Global Outcomes (KDIGO) has also provided additional diagnostic measures for consideration, such as diastolic and systolic blood pressure values less than 90 mmHg and 140 mmHg as positive indicators of chronic kidney disease.^1^

CKD treatment seeks to reduce the progression of the disease by dealing with underlying conditions that accompany the conditions, such as glomerular hypertension, hypertrophy, scarring, and fibrosis. Management strategies geared toward reducing hypertension also allow for the delayed progression of the disease to be effected. As chronic renal failure is associated with increased acidosis, the administration of alkaline solutions, such as sodium bicarbonate, helps to introduce the effects and, therefore, the progression of renal failure.^3^ Glycemic control is also necessary among patients to deal with increased albumin levels. Lifestyle changes are also necessary for people with chronic kidney disease, as increased lipid levels can result in adverse outcomes.^1^ Such changes include dietary management to help reduce the progression of the condition by reducing the intake of foods containing specific compounds.^1^ For example, since patients with CKD have proteinuria, they are advised to consume lower than average protein levels in their diets, which studies have found was associated with a 3 g decline in proteinuria per day. According to KDIGO, daily dietary intake should be restricted to 1.3 g/kg of protein.^2^

Calcium, sodium, phosphorus, and vitamin D levels are constantly altered among individuals with chronic kidney disease and should be routinely monitored and appropriate supplementations provided. Guidelines provided by the National Kidney Foundation state that kidney profiles should include glomerular filtration rates and albumin-to-creatinine levels.^4^ Many factors, such as the toxic effects of the drug and the immunocompromised nature of the patients, hinder the efficient treatment of patients with chronic kidney disease.^2^ Nephrotoxins, for example, in drugs such as nonsteroidal anti-inflammatory drugs, cause contraindications for using drugs within this category for CKD patients. Proton pump inhibitors used in some medical settings are contraindicated because of their potential to cause acute interstitial nephritis and atherosclerosis.^2^ Patients undergoing CKD management must undergo at least one annual check-up for markers of kidney disease progression.^5^

When the Glomerular filtration rate is reduced to less than 15 ml/min/1.73 m^2^, the individual’s kidney loses its normal functioning and most of the time requires outside support to perform everyday functions. However, it is also important to note that while dialysis helps individuals manage normal vital kidney function, these functions are limited by ultrafiltration and diffusion. Dialysis can be performed using one of the three main procedures: peritoneal dialysis, continuous renal replacement therapy, and hemodialysis. Peritoneal dialysis is recommended for younger patients because of its ease of performance. Peritoneal dialysis is only practiced in developing or low-income countries because of the reduced requirements for technical skills and resources, such as electricity and facilities.^5,6^ However, peritoneal dialysis is not commonly practiced due to limited awareness of the limited survival rates, with studies indicating that less than 50% of patients adhere to peritoneal dialysis after 2 years. Failure associated with this technique has also been attributed to the peritoneal membrane being an ineffective long-term ultrafiltration membrane for waste removal, primarily due to increased inflammatory responses and peritoneal infection.^7^

A dialyzer, an external filter, is used in the hemodialysis process and acts by separating waste through a countercurrent blood flow system. The fluid within the dialyzer moved in a different direction from the blood flow. In peritoneal dialysis, which uses the peritoneal membrane as the filter, the semi-permeable membrane helps to separate the material for the passage of waste into the desolate. Therefore, dialysis is based on the principle of diffusion of materials through a semi-permeable membrane. In the counter-current blood flow system during peritoneal dialysis, desolate waste, such as serum creatinine and blood urea nitrogen, into the dialysate with diffusion is affected by the size of the particles, so that larger particles diffuse more slowly. Similar to other interventions for chronic kidney disease, dialysis is associated with several complications such as cardiovascular complications, altered hormonal function, and inflammatory responses. As chronic kidney disease is associated with increased inflammatory responses, renal function is disturbed, resulting in the accumulation of metabolic waste. Dialysis helps reduce metabolic waste, which is necessary for the continued function of the kidney, but at the same time induces immunological responses since the membranes used in dialysis are identified as foreign by the host immune system. The activated immune system results in the accumulation of granulocytes, which increase the release of radical oxygen species, resulting in cellular oxidative stress.^8^ The increased inflammatory response during dialysis is also implicated in altered hormonal function, resulting in reduced levels of thyroid hormones and hypertrophy of the left cardiac ventricle, leading to cardiovascular disease.^8^ Continuous renal replacement therapy combines peritoneal and hemodialysis and is usually applied for 24 h or longer using pump-fuelled venovenous circuits that act as supports for the kidney. Technical skills are required to access the appropriate vessels and to apply the pumps, which will provide a blood supply and a permeable membrane through which the diffusion of waste material will occur. Appropriate Solutions are also required to maintain fluid balance through the membrane. CRRT is commonly indicated in patients with hemodynamic alterations, especially in the intensive care unit. Assist recommendation is due to the mechanism of action of this dialysis approach, which filters out fluid at a slower rate than hemodialysis and peritoneal dialysis so that the hemodynamic effects are less severe.

Despite the slower rate of fluid removal while using CRRT, the cumulative volume of fluid removed after 48 h was higher than that of hemodialysis because of the use of larger volumes of fluid through oral and parental medication, nutrition, and supplementary fluid administration. The development and severity of cerebral edema are also reduced in patients exposed to CRRT because the reduced fluid removal rates also reduce the rate at which the mean arterial pressure and cerebral vasodilation increase. To perform CRRT, facilities and equipment such as the dialysate, replacement fluid blood warmer, anticoagulant filters, and blood purifiers should be present. CRRT can be performed using several techniques, such as continuous venovenous hemofiltration, which involves hydrostatic pressure, and continuous renal vein hemodialysis, which uses diffusion. Convection and diffusion are the basis for the continuous venovenous hemodiafiltration method for CRRT.

### Research Aims and Objectives

In this investigation, we sought to explore the studies’ findings by comparing the mortality rates among the three broad categories of dialysis. Therefore, we aimed to compare the following.

- Mortality rates in CRRT vs. hemodialysis.
- Mortality Rates in CRRT vs. PD.
- Mortality rates in patients with PD vs. HD.

## Methodology

### Study Design

This systematic review and meta-analysis were conducted according to the guidelines of the Cochrane methodology and Preferred Reporting Items for Systematic Reviews and Meta-Analyses (PRISMA). The systematic review process was created to eliminate internal and external sources of invalidity through bias and eliminate outdated and low-quality studies. The PRISMA guidelines were used in the literature search phase of the investigation, development of the inclusion/exclusion criteria using the most appropriate framework, study selection, and design of the analysis protocols.

### Literature Search

We conducted database searches from credible databases, including Google Scholar, ResearchGate, PubMed (MEDLINE), Cochrane Library, and Embase. Database searches were conducted in three phases to cover the three comparisons: CRRT versus HD, PD versus HD, and CRRT versus PD. We also screened through the reference lists of several studies and similar reviews, especially because there have been previous meta-analyses and systematic reviews on some parts of the research objectives of our investigation. We limited the search strings used in the databases to the following keywords: ‘mortalities, fatalities, dialysis, hemodialysis, continuous renal replacement therapy (CRRT), Peritoneal dialysis, and comparative.’ Additionally, the search strings were combined using truncations (Asterix), Boolean operators (AND/OR), and field tags.

### Eligibility Criteria

The eligibility criteria for this study were conducted based on the PECO (Participants, Exposures, Comparators, and Outcomes) framework that would give the study that would be included in the final study selection. Therefore, the participants were patients presenting with acute or chronic kidney failure who required assistance with kidney function. We did not place age limits on the study participants to increase the scope of the studies covering the three comparisons of the dialysis modalities under investigation. However, all the included studies were required to provide detailed inclusion criteria of the participants, such as age limits, the type of kidney disease (acute or chronic), presenting clinical signs by the patients (uremia, albuminuria, oliguria, etc.), and previous interventions conducted. The Exposure characteristics for the participants included in the studies were one of the three modalities for dialysis, such as the CRRT, PD, or HD modes. Since these modes also have other types and techniques for performing, the included studies will be required to provide these specifications. The comparators of the included studies will also be one of the three modes of dialysis under investigation. Lastly, this investigation is interested in the outcomes of mortality rates among patients using dialysis modes; hence, the primary outcome of interest.

This investigation will only include studies published in English with full-text access options. We did not place any limits on the study design.

### Data Extraction

Data from the selected studies will be extracted into a pre-set MS Excel sheet, with primary data points including the author, study design(cross-sectional, randomized controlled trial, comparative study, retrospective or prospective study), study duration or dates(start and end date), participants (number), participant demographics (age(mean ± standard deviation) and gender(%female)), inclusion criteria (underlying disease, chronic or acute kidney disease, presenting signs, age categories, subject consent, etc.), the type of dialysis modalities under investigation, and the mortality rates (% per modality group).

### Statistical Analysis

Review Manager version 5.4 (RevMan 5.4; The Nordic Cochrane Center, The Cochrane Collaboration, 2014) was used to conduct the meta-analysis part of the investigation. A random-effects model was applied to compute the effect size, while the I2 statistic was used to judge heterogeneity. We judged heterogeneity in two categories: 0% (complete consistency) to 100% (complete inconsistency), while reliable heterogeneity of the studies was accepted if it appeared at ≤50%. Statistical significance was recognized only if the p-value was < 0.05. Mortality rate comparisons between CRRT vs. HD, CRRT vs. PD, and HD vs. PD were the three outcomes under analysis.

## Results

### Study Selection

The initial search of the PubMed, Cochrane Library, and EMBASE databases resulted in a total of 609 studies, of which 67 were excluded due to duplication. During the title and abstract screening phase, 337 studies were excluded, including reviews (n=196), case reports and literature reviews (n=66), and 54 irrelevant studies due to the outcomes presented. The final full-text screening excluded 196 studies, and an additional 5 studies were identified from the reference lists of other reviews, bringing the final number of studies identified to 14. Figure 1 shows a PRISMA flowchart of the study selection process.

**Figure 1:**
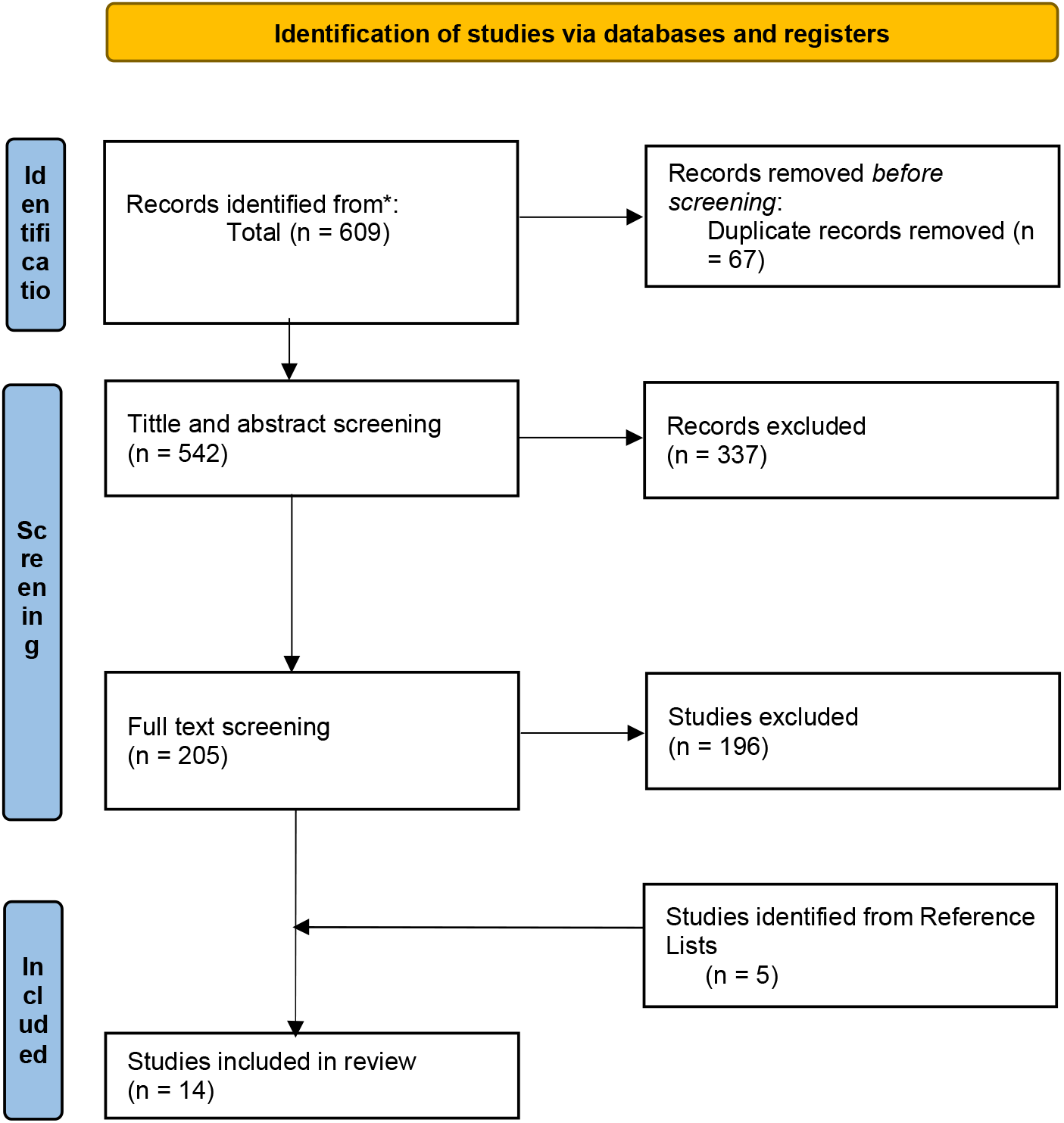
PRISMA flow diagram detailing the study selection process.

**Table 1:** A summary of study characteristics for studies comparing Continuous renal replacement therapy (CRRT) versus Hemodialysis (HD).

| CRRT vs HD |  |  |  |  |  |  |  |  |  |
| --- | --- | --- | --- | --- | --- | --- | --- | --- | --- |
| AUTHOR | STUDY DESIGN | PARTICIPANTS | AGE | GENDER | CRRT | HD | INCLUSION CRITERIA | DURATION | MORTALITY RATES |
| AYDIN, 2022 | Comparative study | 120 | 62.90±13.64 | 38.3% male | 40 | 80 | Patients without end stage kid stage kidney disease | 30 days | CRRT= 67.5%<br>HD = 66.3% |
| GAUNDRY, 2022<sup>9</sup> | Multicentre randomized controlled trial | 543 | Not specified | Not specified | 269 | 274 | Patient informed consent | 60 days | CRRT= 54.3%<br>HD = 46.3% |
| LIANG, 2016<sup>10</sup> | Randomised controlled trial | 638 | 18–44 yrs = 144 (22.6)<br>45–64 yrs = 270 (42.3)<br>65–74 yrs= 128 (20.1)<br>More than 75 yrs. =65 (18.4) 96 (15) | IHD = 57.5% male<br>CRRT= 58.6% | 285 | 353 | Only patients without a history of chronic kidney dialysis or transplant, and creatinine level >4mg/dl and no history of heart failure | 90 days | IHD= 55.9%<br>CRRT = 60% |
| TRUCE, 2019<sup>11</sup> | Prospective observational multicentre cohort | 1360 | 65 (54–76) | 876 (64.4) | 544 | 816 | No history of previous kidney transplant or chronic kidney disease requiring renal replacement therapy | 30 days | CRRT= 46.5%,<br>HD = 35% |

|  |  |  |  |  |  |  |  |  |  |
| --- | --- | --- | --- | --- | --- | --- | --- | --- | --- |
| SCHEFOLD,<br>2014<sup>12</sup> | Single-center | 252 | IHD =60.8 ± 13.4 | IHD = 81 | 122 | 128 | > 18 yrs. Old, patients | 30 days | CRRT =45.4% |
|  | prospective |  | CRRT = 62.3 ± 14.5 | (63.3% |  |  | requiring RRT, clinical |  | HD = 52.4% |
|  | randomized |  |  | female) |  |  | signs of uraemia, oliguria, |  |  |
|  | controlled trial |  |  | CRRT = |  |  | anuria, and severe |  |  |
|  |  |  |  | 75 (61.5% |  |  | metabolic acidosis |  |  |
|  |  |  |  | male) |  |  |  |  |  |

**Table 2:**
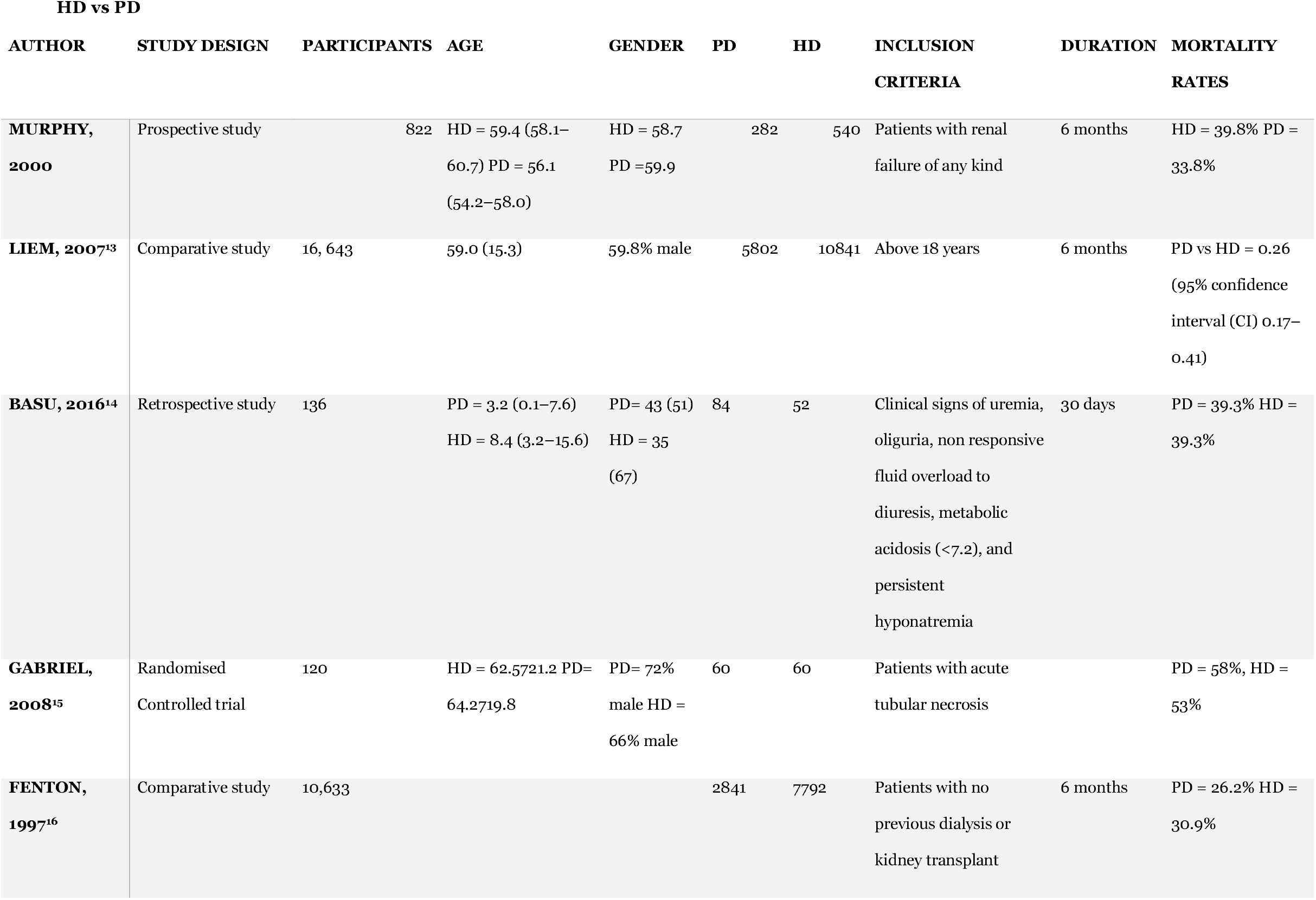
A summary of study characteristics for studies comparing Hemodialysis (HD) versus Peritoneal Dialysis (PD).

**Table 3:** A summary of study characteristics for studies comparing Peritoneal Dialysis (PD) versus Continuous renal replacement therapy (CRRT).

| PD vs CRRT |  |  |  |  |  |  |  |  |  |
| --- | --- | --- | --- | --- | --- | --- | --- | --- | --- |
| AUTHOR | STUDY DESIGN | PARTICIPANTS | AGE | GENDER | CRRT | PD | INCLUSION CRITERIA | DURATION | MORTALITY RATES |
| PHU, 2002<sup>17</sup> | Randomised controlled Trial | 70 | PD = 36 (29.6–38.4) CRRT= 35 (29.5–38.2 | PD = 75% male, CRRT = 88% male | 34 | 36 | Written and informed consent. Above 15 years, without previous history of RRT | 1 year | PD = 47% CRRT= 15% |
| JAYRAL, 2017<sup>18</sup> | Retrospective analysis deisgn | 40 | 53.5 + 17 | 57.5% male | 22 | 18 | More than 12 yrs., and glomerular filtration rate <60 ml/min/1.73 m2 | 1 year | CRRT =81.8% PD = 77.8% |
| AL-HWIESH, 2018<sup>19</sup> | Prospective Randomized Study | 125 | CRRT = 44.60 12.38 PD = 45.41 14.12 | CRRT= 72.6% PD= 74.6% | 62 | 63 | Written and informed consent. Above 18 years | 28 days | 53.2% = CRRT PD= 30.2% |
| GEORGE, 2011<sup>20</sup> | Open prospective randomized study | 55 | CRRT = 45.32±17.53 yrs, PD= 45.32±17.53 yrs. | CRRT = 60% PD = 64% | 25 | 25 | Patients who did not undergo previous abdominal surgery or without any pulmonary edema | 3 yrs. | CRRT = 84% PD = 72% |

### Risk of Bias

**Figure 2:**
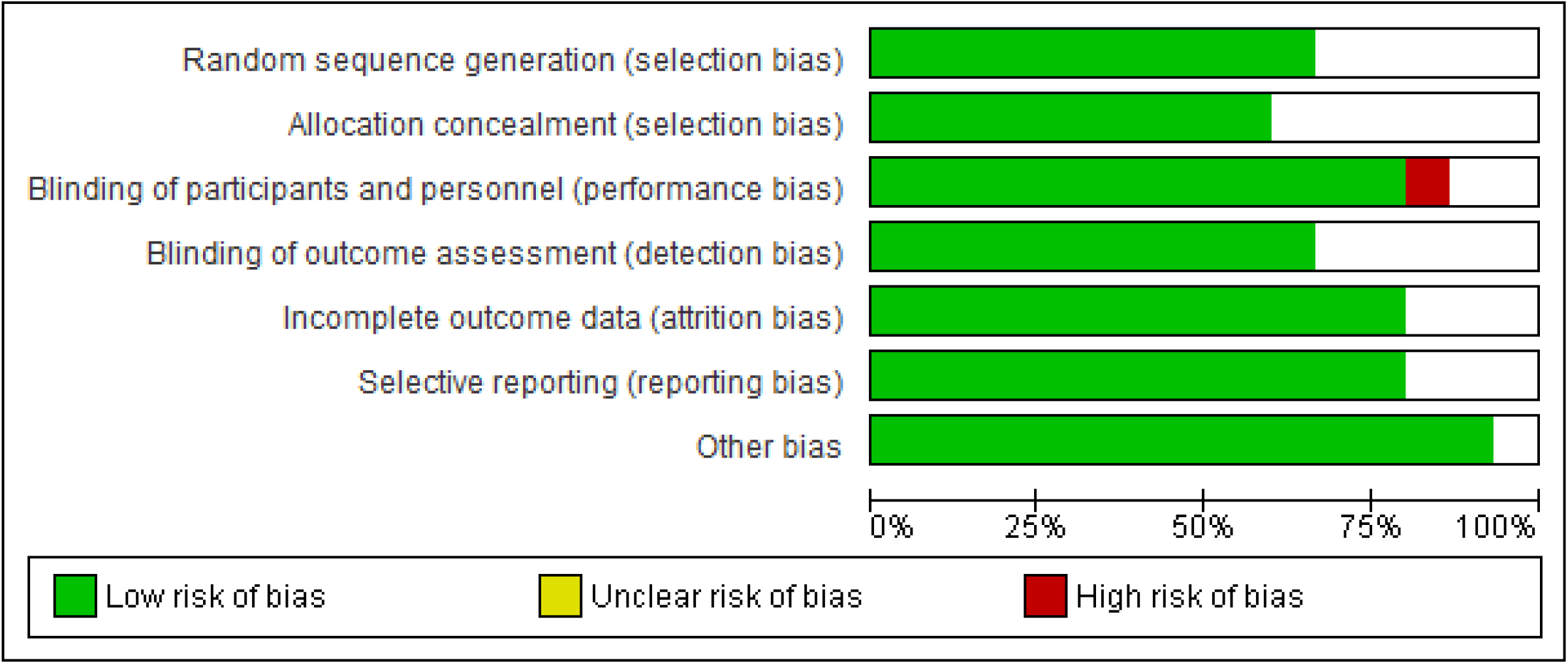
Risk of bias graph: Review authors’ judgments about each risk of bias item presented as percentages across all included studies.

**Figure 3:**
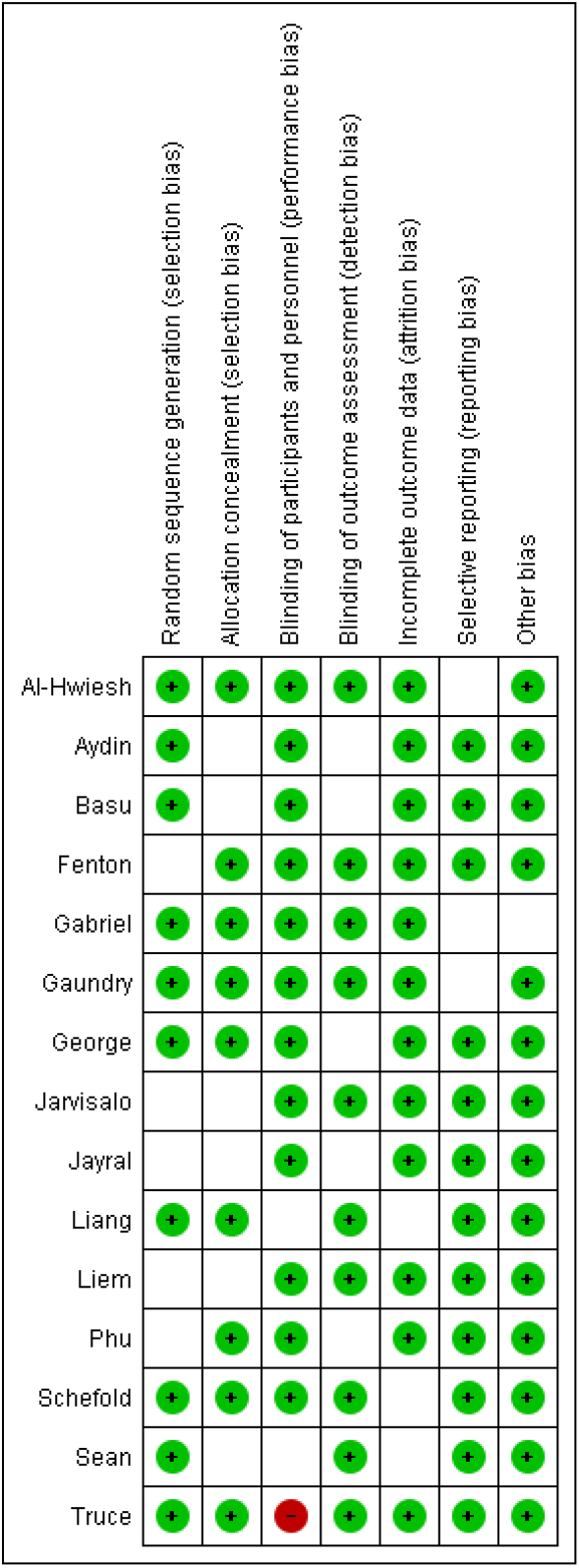
Risk of bias summary: review authors’ judgments about each risk of bias item for each included study

### Meta-Analysis

#### CRRT vs. PD

Four of the selected studies analyzed the mortality rates after using CRRT and PD dialysis modalities.^18-20^. A total of 143 and 142 patients were categorized into the CRRT and PD groups, respectively. The random effects risk ratio was 0.98 [0.62, 1.54] at a 95% confidence interval. No statistically significant difference was found in mortality rates between the two randomized groups (p= 0.92). The included studies had a high level of heterogeneity (p= 0.02, I^2^ = 71%).

#### PD vs. HD

Four selected studies analyzed the mortality rates after HD and PD dialysis modalities^14-16,21^ A total of 3267 patients were categorized into the PD group, whereas 8444 were categorized into the HD group. The random effects risk ratio was 0.83 [0.59, 0.87] at a 95% confidence interval. No statistically significant difference was found in mortality rates between the two randomized groups (p= 0.09). The included studies had a high level of heterogeneity (p< 0.00002, I ^2^ = 90%). Figure 6 shows a forest plot of the above results, with funnel plots (figure 7) showing the levels of publication bias among the selected studies.

**Figure 4:**
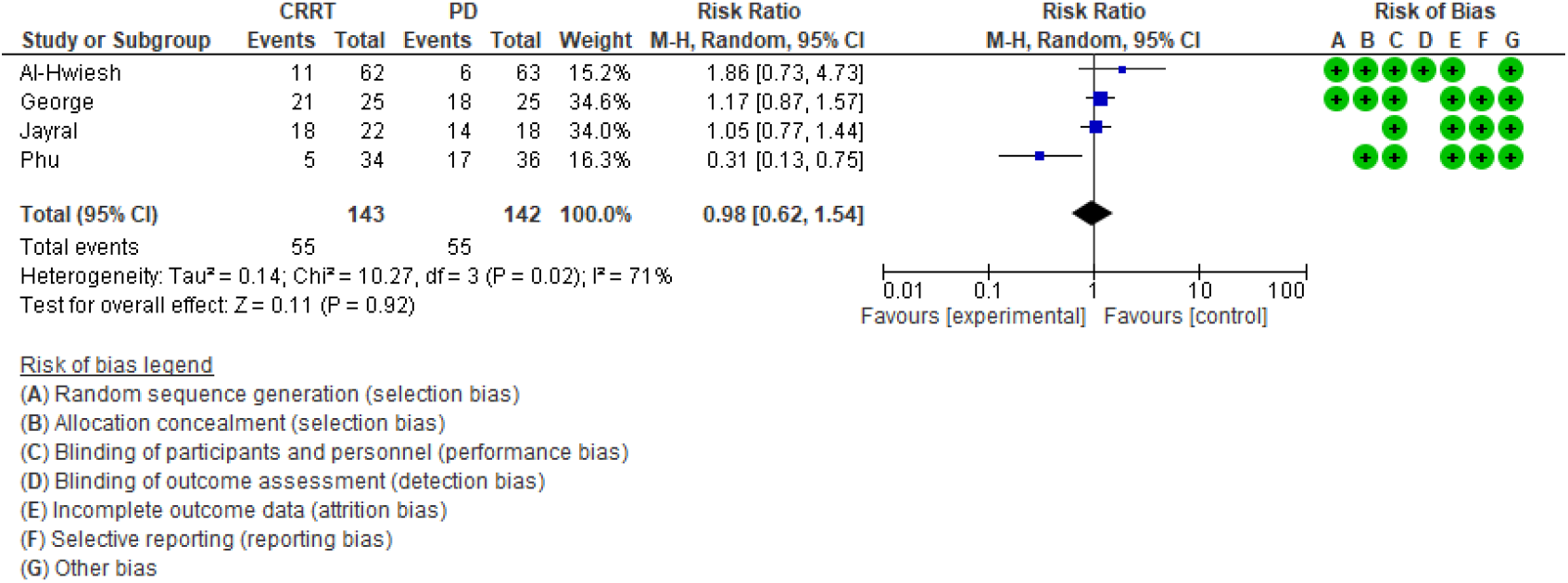
A forest plot for the incidence of mortality between Continuous renal replacement therapy (CRRT) and Peritoneal Dialysis (PD).

**Figure 5:**
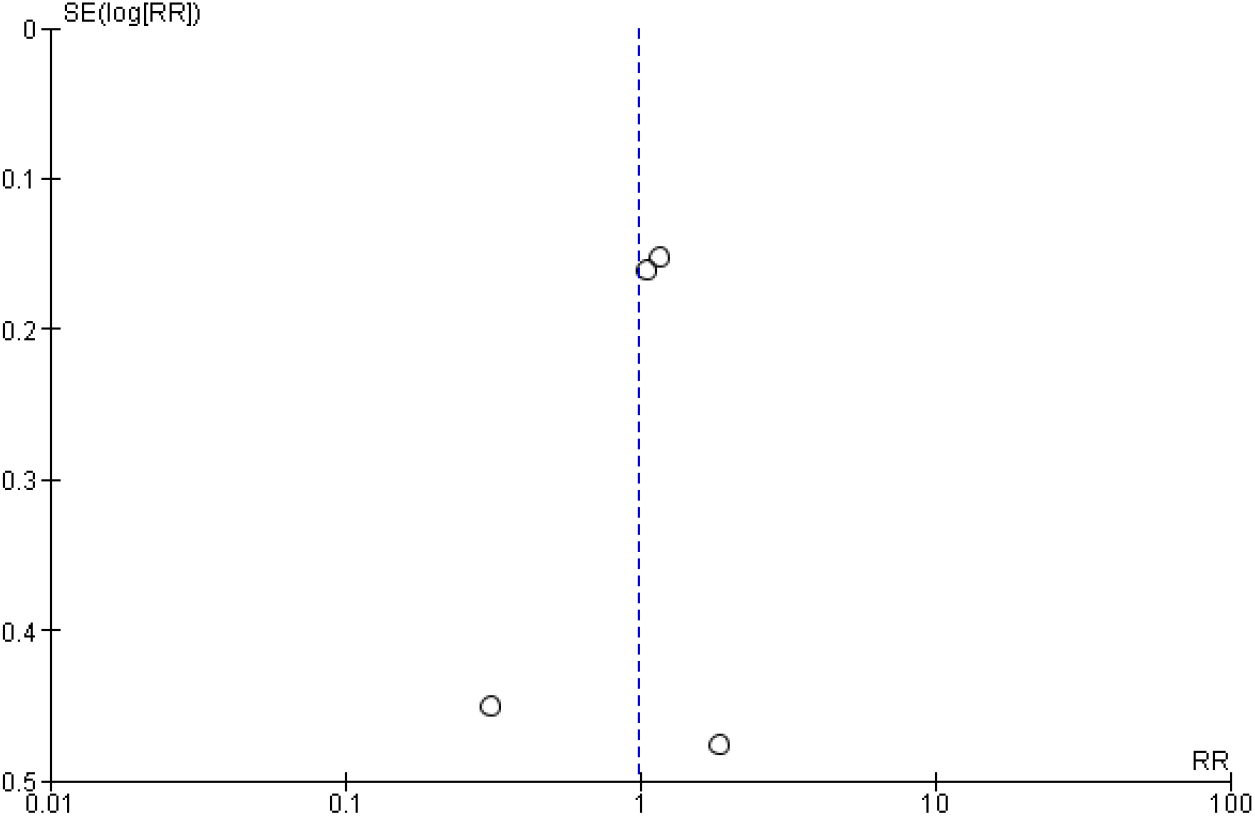
Funnel plot representing the publication bias between the selected studies.

**Figure 6:**
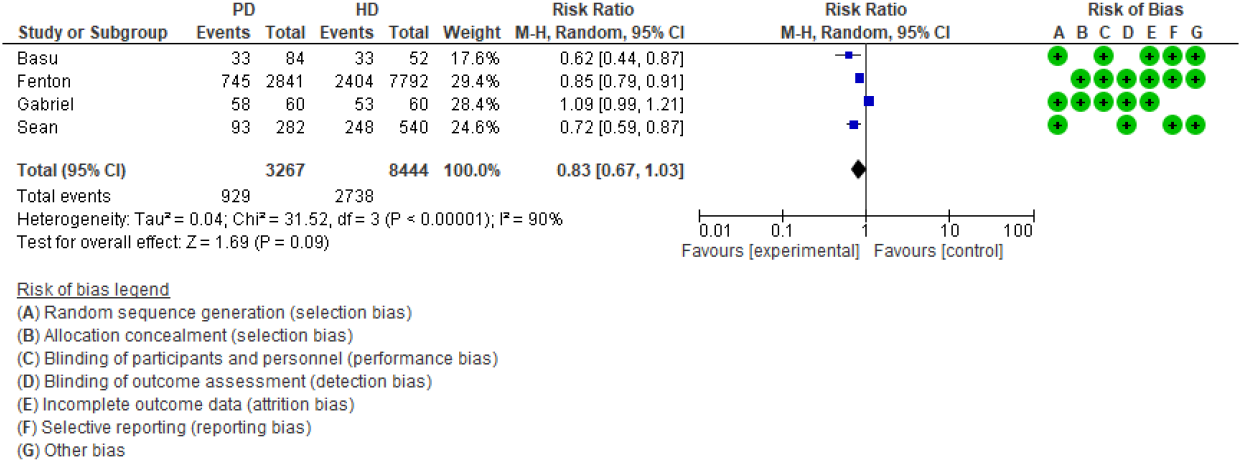
A forest plot for the incidence of mortality between Peritoneal Dialysis (PD) and Hemodialysis (HD).

**Figure 7:**
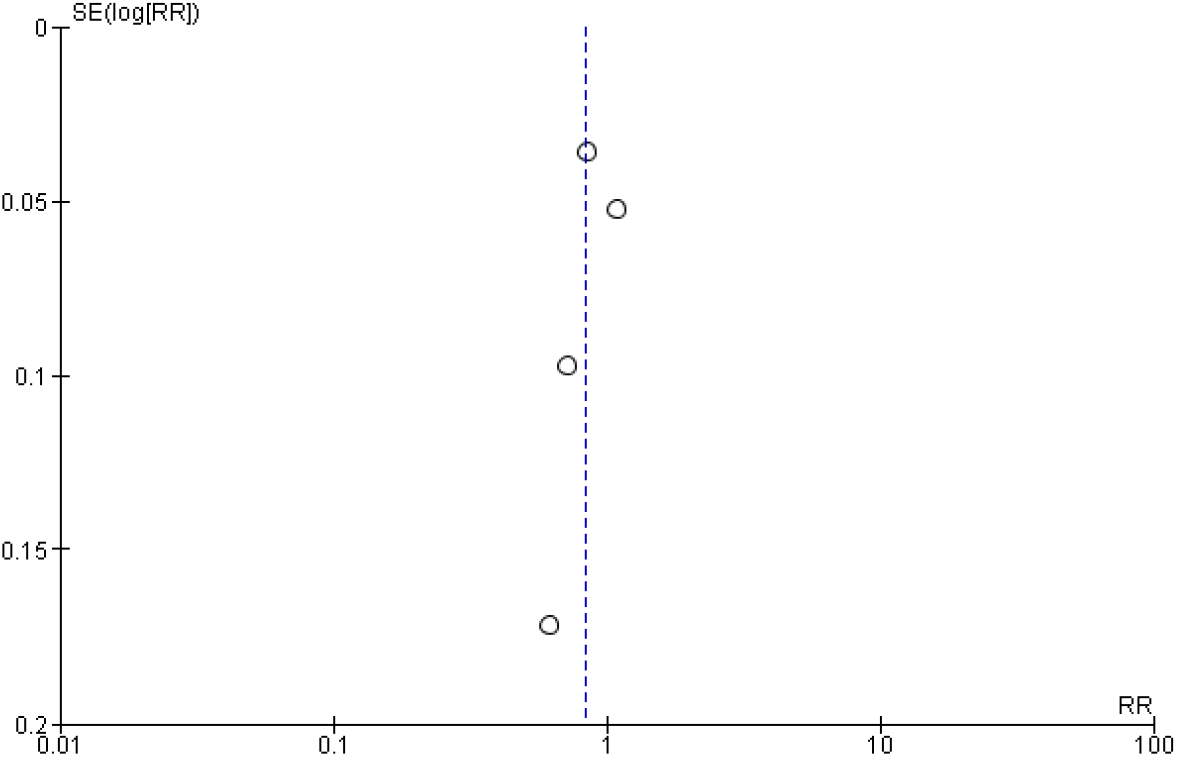
Funnel plot representing the publication bias between the selected studies.

#### HD vs. CRRT

Five of the selected studies analyzed the mortality rates after CRRT and HD dialysis modalities.^9-12,22^ A total of 1045 patients were categorized into the CRRT group, while 1416 were in the HD group. The random effects risk ratio was 1.10 [0.95, 1.27] at a 95% confidence interval. No statistically significant difference was found in mortality rates between the two randomized groups (p= 0.22). The included studies had a moderately high level of heterogeneity (p= 0.03, I ^2^ = 63%).

**Figure 8:**
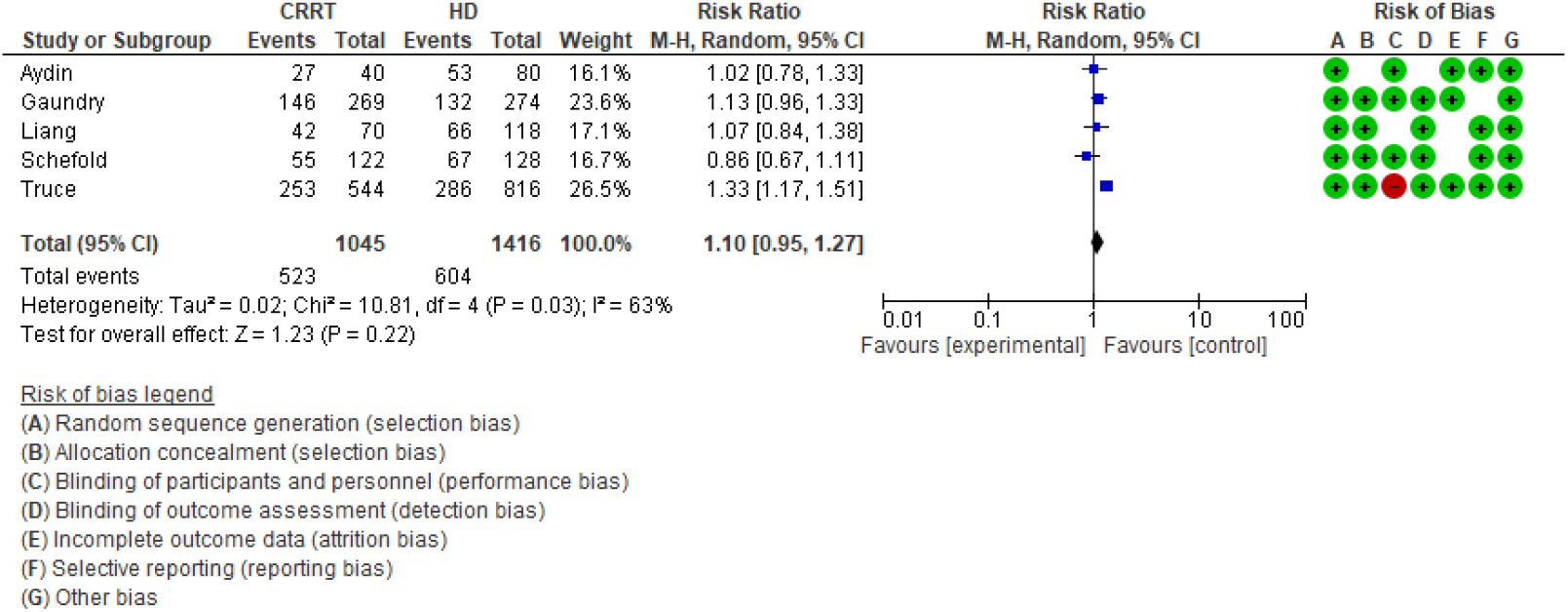
A forest plot for the incidence of mortality between Continuous renal replacement therapy (CRRT) and Hemodialysis (HD).

**Figure 9:**
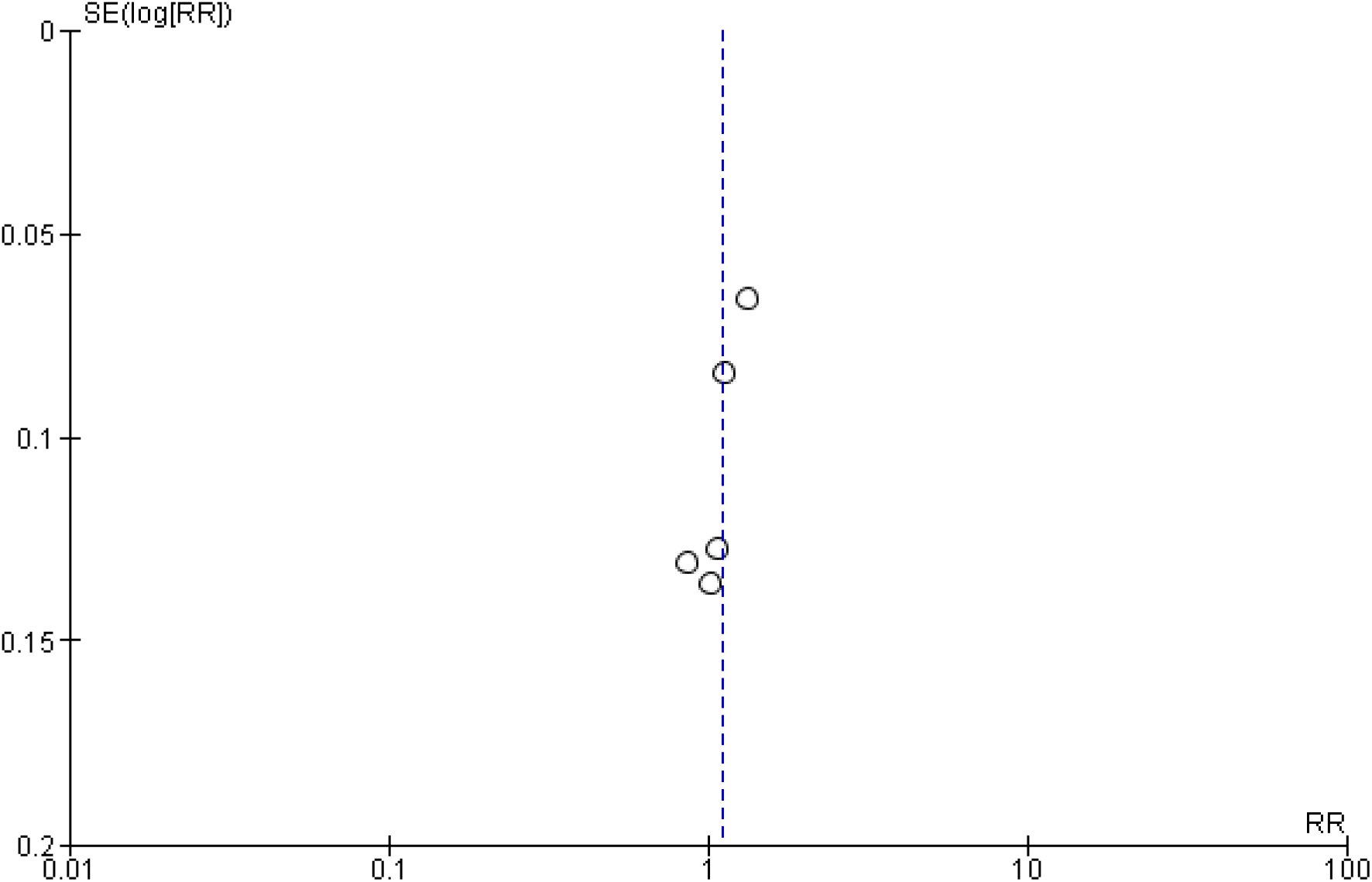
Funnel plot representing the publication bias between the selected studies.

## Discussion

### CRRT vs. PD

This investigation found no statistically significant differences in mortality among patients receiving CRRT and PD dialysis (p= 0.92). These results are from the results of the meta-analysis, which correspond to the study by George et al., who reported that while there were higher mortality rates among patients receiving CRRT than PD dialysis mortalities, the differences between the two were not statistically significant (84% vs. 72%, p=0.49).^20^ However, the study by Al-Hwiesh et al. reported similar findings in terms of mortality rates but conflicting findings in terms of statistical significance (p=0.0028).^19^ Similar to the study by Al-Hwiesh et al., Phu et al. reported a 47% mortality rate in the PD group, while 15% was recorded in the CRRT group (p=0.005).^19^

### PD vs. HD

Similarly, the difference in the occurrence of mortality between patients receiving PD and HD did not reflect any statistical significance (p = 0.09). These findings were supported by those of the included studies, such as that by Gabriel et al., who reported a 58% mortality rate among patients receiving PD compared to the 53% rate among those receiving HD; however, the difference was not statistically significant (p= 0.48).^15^ Fenton et al. also reported higher mortality rates among patients receiving hemodialysis than among those receiving HD, but the difference was not statistically significant. The mortality rate also seemed to vary according to the age groups reported by the study, with all age groups indicating higher rates of mortality among patients receiving hemodialysis than among those receiving peritoneal dialysis.^16^ At 6 months follow-up, Murphy et al. reported higher mortality rates among patients receiving HD than among those receiving PD therapy. Worsening cardiovascular symptoms, age, and diabetes are commonly associated with mortality risk factors among patients.^21^

### HD vs. CRRT

The difference in the mortality rates between patients receiving HD and CRRT therapy (p= 0.22). The lack of statistical significance was also noted in the study by Liang et al., who reported higher death rates among patients receiving CRRT modalities than those in the HD therapy group within the 90-day study period (0.65).^10^ Using the Kaplan-Meier Scale, the death rate was reported to be higher in the CRRT group than in the HD group within the 60-day study duration (HR 1.27, 95% CI 1.00 to 1.61).^9^ Aydin et al. noted no statistical significance in the differences in mortality rates among patients in the HD and CRRT groups (66.3% vs. 67.5%, p= 0.891). ^22^

## Conclusion

This investigation sought to explore the differences in the mortality rates among patients receiving different modes of dialysis (PD, CRRT, or HD). We found that mortality resulting from various factors, including underlying systemic diseases such as hypertension, diabetes, and hypothyroidism, usually worsened during medical interventions, drugs, and even during dialysis. We found no statistically significant differences between the three groups in the occurrence of mortality among the patients.

The systematic review and meta-analysis were conducted to include different techniques of the three primary dialysis modalities. Therefore, we did not limit the search to specific techniques of CRRRT, HD, or PD; hence recommend that future studies aim to explore the efficiency of the dialysis modalities.

## Data Availability

All data produced in the present work are contained in the manuscript

Endnote References.

## References

1. Chen TK, Knicely DH, Grams ME. Chronic Kidney Disease Diagnosis and Management: A Review. JAMA 2019;322(13):1294–1304. DOI: 10.1001/jama.2019.14745.

2. Vadakedath S, Kandi V. Dialysis: A Review of the Mechanisms Underlying Complications in the Management of Chronic Renal Failure. Cureus 2017;9(8):e1603. DOI: 10.7759/cureus.1603.

3. Kim HJ. Metabolic Acidosis in Chronic Kidney Disease: Pathogenesis, Clinical Consequences, and Treatment. Electrolyte Blood Press 2021;19(2):29–37. DOI: 10.5049/EBP.2021.19.2.29.

4. Grams ME, Astor BC, Bash LD, Matsushita K, Wang Y, Coresh J. Albuminuria and estimated glomerular filtration rate independently associate with acute kidney injury. J Am Soc Nephrol 2010;21(10):1757–64. DOI: 10.1681/ASN.2010010128.

5. Diamantidis CJ, Hale SL, Wang V, Smith VA, Scholle SH, Maciejewski ML. Lab-based and diagnosis-based chronic kidney disease recognition and staging concordance. BMC Nephrol 2019;20(1):357. DOI: 10.1186/s12882-019-1551-3.

6. Zimmerman AM. Peritoneal dialysis: increasing global utilization as an option for renal replacement therapy. J Glob Health 2019;9(2):020316. DOI: 10.7189/jogh.09.020316.

7. Witowski J, Lopez-Cabrera M. Peritoneal Dialysis and Its Local and Systemic Complications: From the Bench to the Clinic. Front Physiol 2020;11:188. DOI: 10.3389/fphys.2020.00188.

8. Dabrowska-Bender M, Dykowska G, Zuk W, Milewska M, Staniszewska A. The impact on quality of life of dialysis patients with renal insufficiency. Patient Prefer Adherence 2018;12:577–583. DOI: 10.2147/PPA.S156356.

9. Gaudry S, Grolleau F, Barbar S, et al. Continuous renal replacement therapy versus intermittent hemodialysis as first modality for renal replacement therapy in severe acute kidney injury: a secondary analysis of AKIKI and IDEAL-ICU studies. Crit Care 2022;26(1):93. DOI: 10.1186/s13054-022-03955-9.

10. Liang KV, Sileanu FE, Clermont G, et al. Modality of RRT and Recovery of Kidney Function after AKI in Patients Surviving to Hospital Discharge. Clin J Am Soc Nephrol 2016;11(1):30–8. DOI: 10.2215/CJN.01290215.

11. Truche AS, Darmon M, Bailly S, et al. Erratum to: Continuous renal replacement therapy versus intermittent hemodialysis in intensive care patients: impact on mortality and renal recovery. Intensive Care Med 2016;42(9):1523. DOI: 10.1007/s00134-016-4418-0.

12. Schefold JC, von Haehling S, Pschowski R, et al. The effect of continuous versus intermittent renal replacement therapy on the outcome of critically ill patients with acute renal failure (CONVINT): a prospective randomized controlled trial. Crit Care 2014;18(1):R11. DOI: 10.1186/cc13188.

13. Liem YS, Wong JB, Hunink MG, de Charro FT, Winkelmayer WC. Comparison of hemodialysis and peritoneal dialysis survival in The Netherlands. Kidney Int 2007;71(2):153–8. DOI: 10.1038/sj.ki.5002014.

14. Basu B, Mahapatra TK, Roy B, Schaefer F. Efficacy and outcomes of continuous peritoneal dialysis versus daily intermittent hemodialysis in pediatric acute kidney injury. Pediatr Nephrol 2016;31(10):1681–9. DOI: 10.1007/s00467-016-3412-7.

15. Gabriel DP, Caramori JT, Martim LC, Barretti P, Balbi AL. High volume peritoneal dialysis vs daily hemodialysis: a randomized, controlled trial in patients with acute kidney injury. Kidney Int Suppl 2008(108):S87–93. DOI: 10.1038/sj.ki.5002608.

16. Fenton SS, Schaubel DE, Desmeules M, et al. Hemodialysis versus peritoneal dialysis: a comparison of adjusted mortality rates. Am J Kidney Dis 1997;30(3):334–42. DOI: 10.1016/s0272-6386(97)90276-6.

17. Phu NH, Hien TT, Mai NT, et al. Hemofiltration and peritoneal dialysis in infection-associated acute renal failure in Vietnam. N Engl J Med 2002;347(12):895–902. DOI: 10.1056/NEJMoa020074.

18. Jaryal A, Vikrant S. A Study of Continuous Renal Replacement Therapy and Acute Peritoneal Dialysis in Hemodynamic Unstable Patients. Indian J Crit Care Med 2017;21(6):346–349. DOI: 10.4103/ijccm.IJCCM_143_17.

19. Al-Hwiesh A, Abdul-Rahman I, Finkelstein F, et al. Acute Kidney Injury in Critically Ill Patients: A Prospective Randomized Study of Tidal Peritoneal Dialysis Versus Continuous Renal Replacement Therapy. Ther Apher Dial 2018;22(4):371–379. DOI: 10.1111/1744-9987.12660.

20. George J, Varma S, Kumar S, Thomas J, Gopi S, Pisharody R. Comparing continuous venovenous hemodiafiltration and peritoneal dialysis in critically ill patients with acute kidney injury: a pilot study. Perit Dial Int 2011;31(4):422–9. DOI: 10.3747/pdi.2009.00231.

21. Murphy SW, Foley RN, Barrett BJ, et al. Comparative mortality of hemodialysis and peritoneal dialysis in Canada. Kidney Int 2000;57(4):1720–6. DOI: 10.1046/j.1523-1755.2000.00017.x.

22. Yilmaz Aydin F, Aydin E, Kadiroglu AK. Comparison of the Treatment Efficacy of Continuous Renal Replacement Therapy and Intermittent Hemodialysis in Patients With Acute Kidney Injury Admitted to the Intensive Care Unit. Cureus 2022;14(1):e21707. DOI: 10.7759/cureus.21707.

